# seg-metrics: a Python package to compute segmentation metrics

**DOI:** 10.1101/2024.02.22.24303215

**Authors:** Jingnan Jia, Marius Staring, Berend C. Stoel

## Abstract

Medical image segmentation (MIS) is an important task in medical image processing. Unfortunately, there is not a out-of-the-box python package for the evaluation metrics of MIS. Therefore, we developed `seg-metrics`, an open-source Python package for MIS model evaluation. Unlike existing packages, `seg-metrics` offers user-friendly interfaces for various overlap-based and distance-based metrics, providing a comprehensive solution. `seg-metrics` supports multiple file formats and is easily installable through the Python Package Index (PyPI). With a focus on speed and convenience, `seg-metrics` stands as a valuable tool for efficient MIS model assessment.

## 1 Background

In the last decade, the research of artificial intelligence on medical images has attracted researchers’ interest. One of the most popular directions is automated medical image segmentation (MIS) using deep learning, which aims to automatically assign labels to pixels so that the pixels with the same label from a segmented object. However, in the past years a strong trend of highlighting or cherry-picking improper metrics to show particularly high scores close to 100% was revealed in scientific publishing of MIS studies [1]. In addition, even though there are some papers that evaluate image segmentation results from different perspectives, the implementation of their evaluation algorithms is inconsistent. This is due to the lack of a universal metric library in Python for standardized and reproducible evaluation. Therefore, we proposed to develop an open-source publicly available Python package `seg-metrics`, which aims to evaluate the performance of MIS models. Our package is public available at https://pypi.org/project/seg-metrics.

## 2 Related packages

As far as we know, untill the publication date of this package (2020), there are only two open source packages which could perform MIS metrics calculation: `SimpleITK`[2] and `Medpy` [3].

**SimpleITK** is an interface (including Python, c#, Java, and R) to the Insight Segmentation and Registration Toolkit (ITK) designed for biomedical image analysis. Unfortunately, `SimpleITK` does not support the evaluation of MIS directly. Each evaluation consists of several basic steps, which makes it not user-friendly. **Medpy** is a medical image processing library written in Python. It includes some functions to evaluate MIS. However, it mainly support the operations of binary segmentation results, which limits its wider application scenarios. Therefore, this work aims to develop a Python package specifically for MIS.

## 3 Our seg-metrics package

Our `seg-metrics` package supports calculating different evaluation metrics directly in one line of code. The metrics could be divided to overlap-based metrics and distance-based metrics. Overlap-based metrics, define the overlap between the reference annotation and the prediction of the algorithm. It is typically complemented by a distance-based metrics, which could explicitly assess how close the boundaries are between the prediction and the reference [4]. The details of the two categories are described below.

### 3.1 Overlap-based metrics

A confusion matrix (see Table 1) could be derived when comparing a segmentation (pixelwise classification) result and its reference. In this table, there are 4 different outcomes:

**Table 1:**
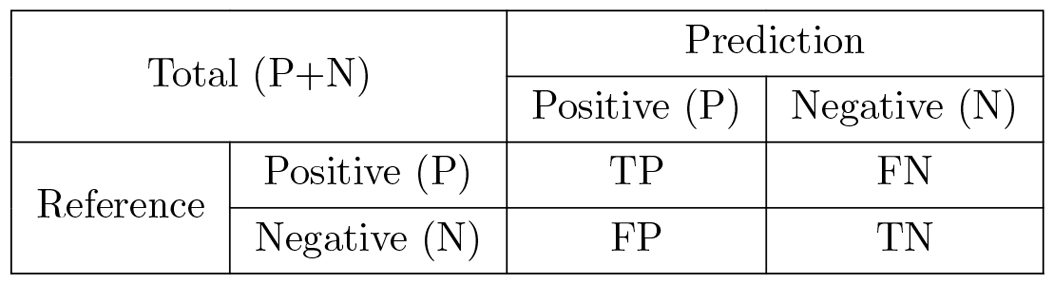
Confusion matrix (adopted from https://en.wikipedia.org/wiki/Confusion_matrix)

| Total (P+N) |  | Prediction |  |
| --- | --- | --- | --- |
|  |  | Positive (P) | Negative (N) |
| Reference | Positive (P) | TP | FN |
|  | Negative (N) | FP | TN |

1. **TP**: If the actual classification is positive and the predicted classification is positive, this is called a true positive (TP) result because the positive sample was predicted correctly.
2. **FN**: If the actual classification is positive and the predicted classification is negative, this is called a false negative (FN) result because the positive sample is incorrectly predicted as being negative.
3. **FP**: If the actual classification is negative and the predicted classification is positive, this is called a false positive (FP) result because the negative sample is incorrectly predicted as being positive.
4. **TN**: If the actual classification is negative and the predicted classification is negative, this is called a true negative (TN) result because the negative sample is predicted correctly.

Based on these four outcomes, we can derive a great number of overlap-based metrics. Their equations are as follows.

- **Dice Coefficient (F1-Score)**

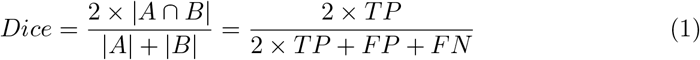
- **Jaccard index**

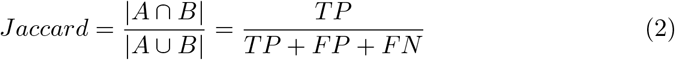
- **Precision/Positive predictive value (PPV)** Precision score is the number of true positive results divided by the number of all positive results

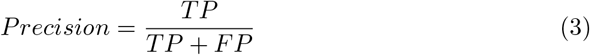
- **Selectivity/Specificity/True negative rate (TNR)**

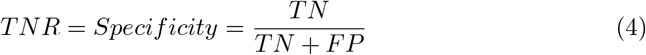
- **False negative rate (FNR)**

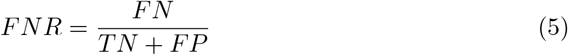
- **Recall/Sensitivity/Hit rate/True positive rate (TPR)** Recall score, also known as Sensitivity, hit rate, or TPR, is the number of true positive results divided by the number of all samples that should have been identified as positive

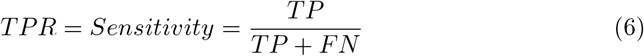
- **False positive rate (FPR)**

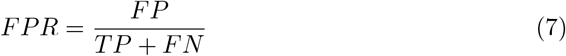
- **Accuracy/Rand Index** Accuracy score, also known as Rand index is the number of correct predictions, consisting of correct positive and negative predictions divided by the total number of predictions.

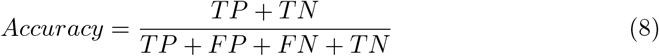
- **Volume similarity** Volume similarity measures the absolute size difference of the regions, as a fraction of the size of the sum of reference and segmentation result. There is more than one definations for the volume similarity [5].
  1. The first definition is [5]:

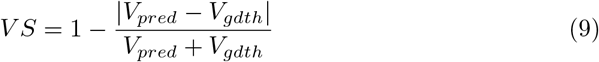

where *V*_*pred*_ is the volume of prediction and *V*_*gdth*_ is the volume of the ground truth. It ranges from 0 to 1. Higher value means the size (volume) of the prediction is more similar (close) with the size (volume) of the ground truth.
  2. The second definition is:

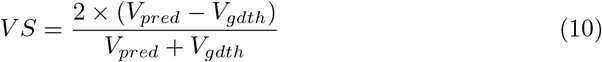

This definition is from the official tutorial of `SimpleITK` [6]. Negative VS means the volume of prediction is less than the volume of ground truth, which is called **underestimation**. Positive VS means the volume of prediction is greater than the volume of the ground truth, which is called **overestimation**.

In our package `seg_metrics`, we implemented the **second** definition. Please note that none of the two equations represent overlap information. VS only represents the volume size difference between prediction and ground truth.

### 3.2 Distance-based metrics

- **Hausdorff distance (HD)** (see Figure 1)

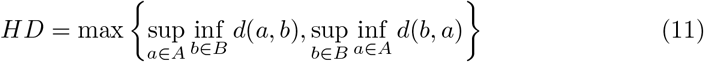

where *sup* represents the supremum operator, *inf* is the infimum operator, and *inf*_*b*∈*B*_*d*(*a, b*) quantifies the distance from a point *a* ∈ *X* to the subset *B* ⊆ *X*.
- **Hausdorff distance 95% percentile (HD95)** is the 95% percentile of surface distances between segmentation and reference.
- **Mean (Average) surface distance (MSD)** is the mean value of surface distances between segmentation and reference [7, 8].
- **Median surface distance (MDSD)** is the median value of surface distances between segmentation and reference. **Note:** These metrics are **symmetric**, which means the distance from segmentation result to reference is the same as the distance from reference to segmentation result.

## 4 Installation

Our package was published in the Python Package Index (PyPI), which is the official third-party software repository for Python. Thus, `seg-metrics` can be directly installed and immediately used in any Python environment using a single line as follows.

**Figure 1:**
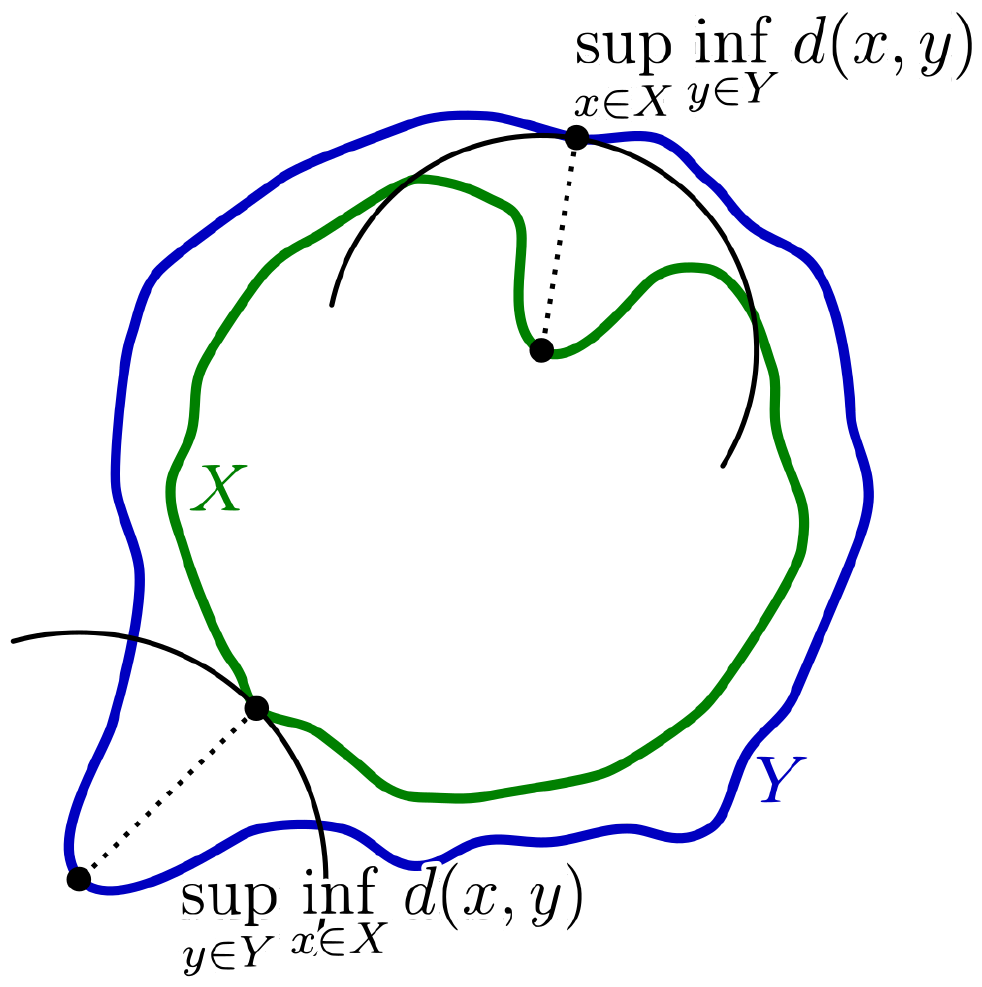
Hausdorff distance between the green curve X and the blue curve Y.

~~~
$ pip install seg-metrics
~~~

## 5 Use cases

`seg-metrics` is a Python package which outputs the segmentation metrics by receiving one ground truth image and another predicted image. After we import the package by “`from seg metrics import seg metrics`”, the syntax to use it is as follow (**Note:** all the following cases are based on textttseg-metrics 1.1.6).

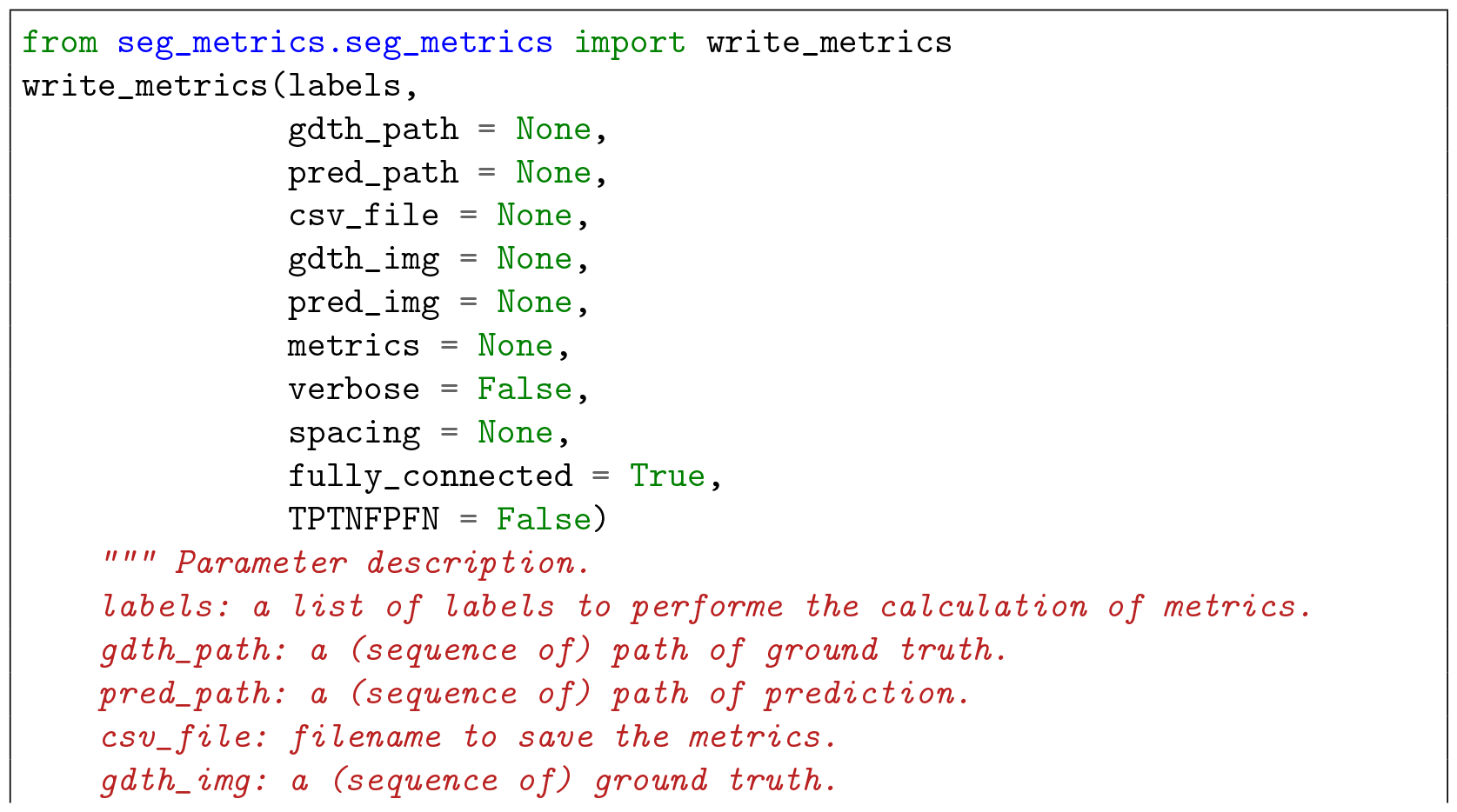
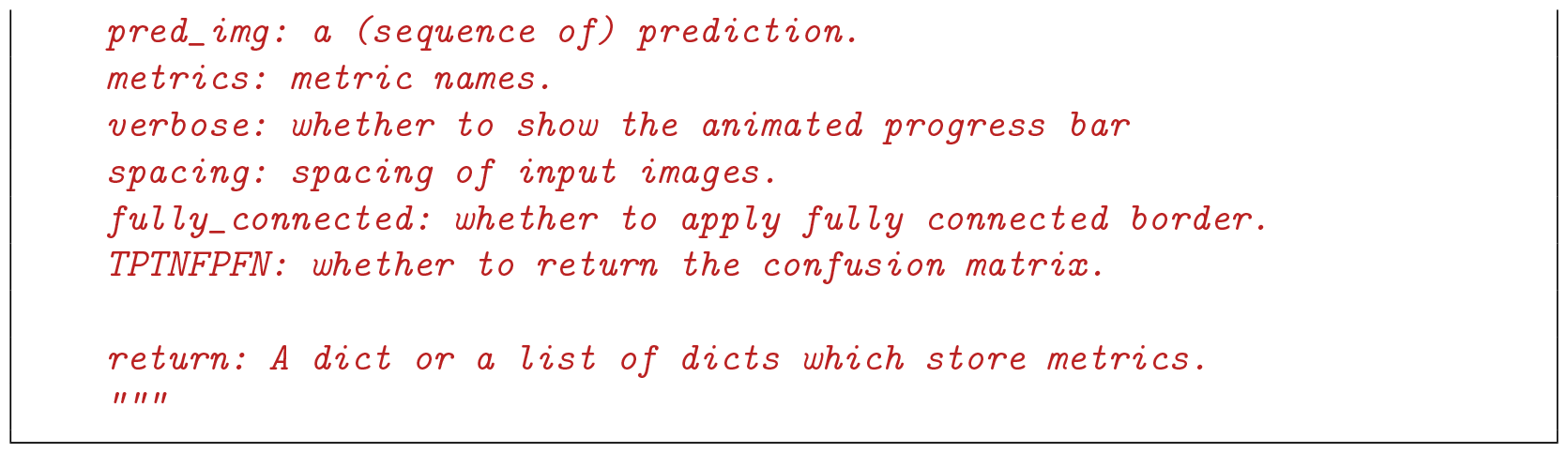

More examples are shown below.

- Evaluate two batches of images with same filenames from two different folders.

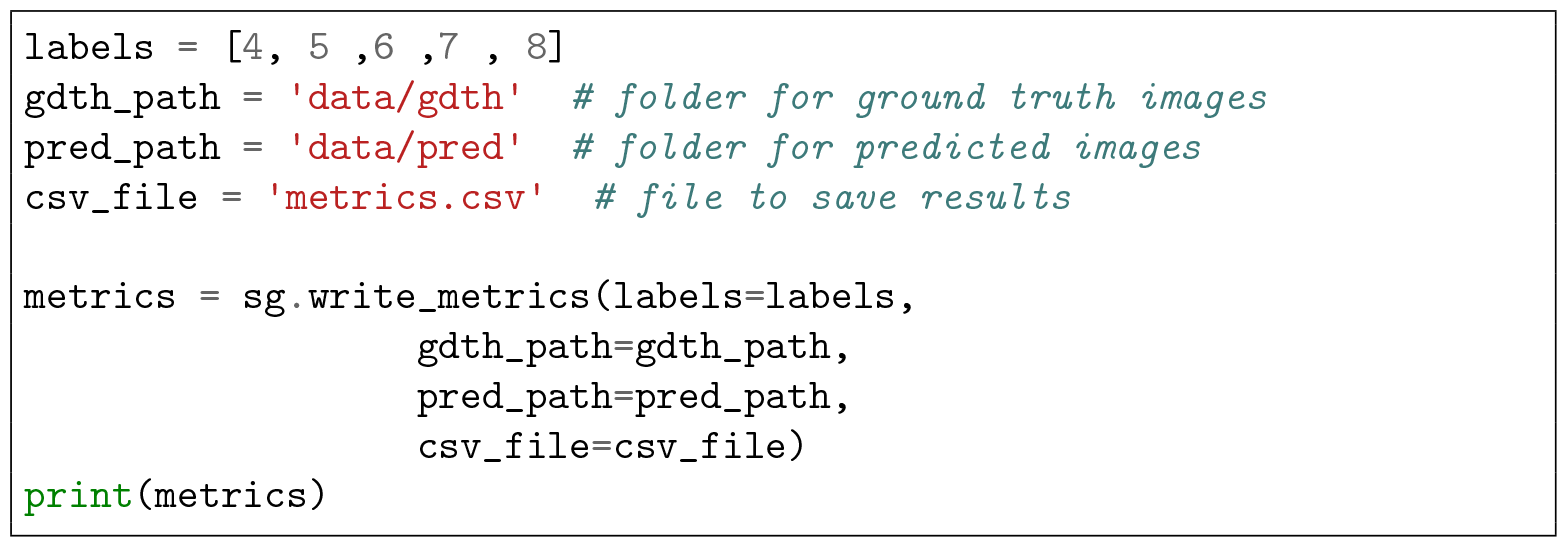
- Evaluate two images

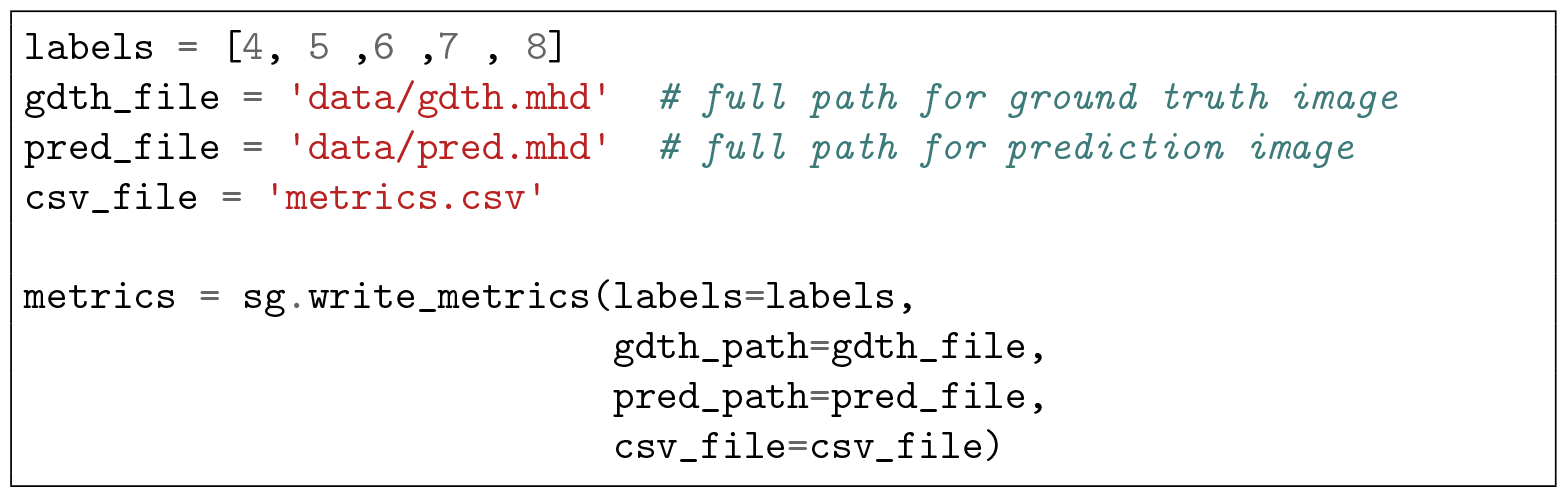
- Evaluate two images with specific metrics

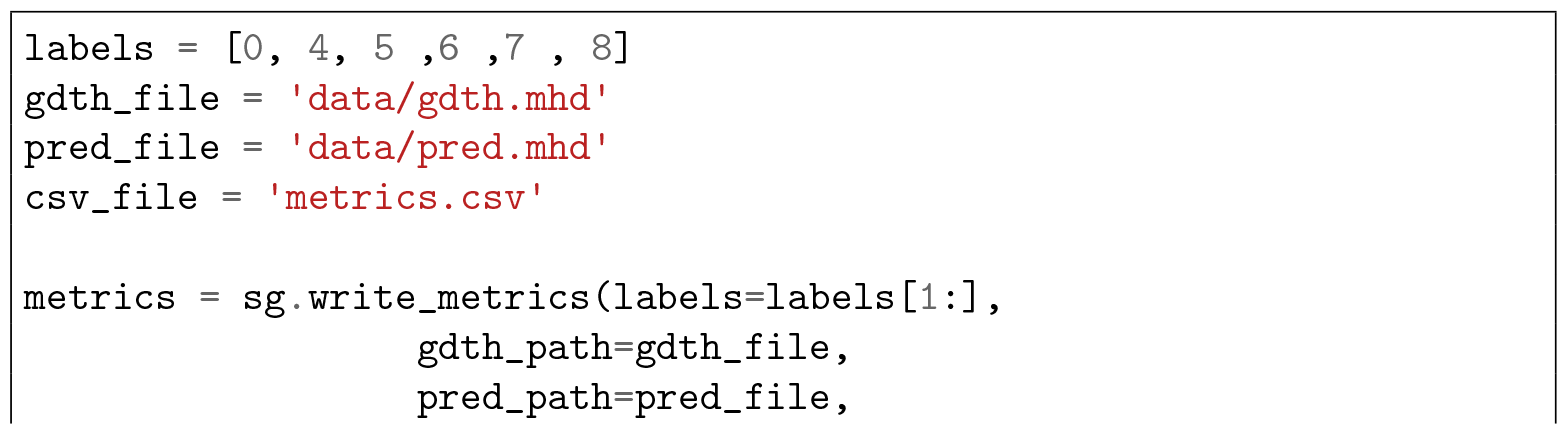
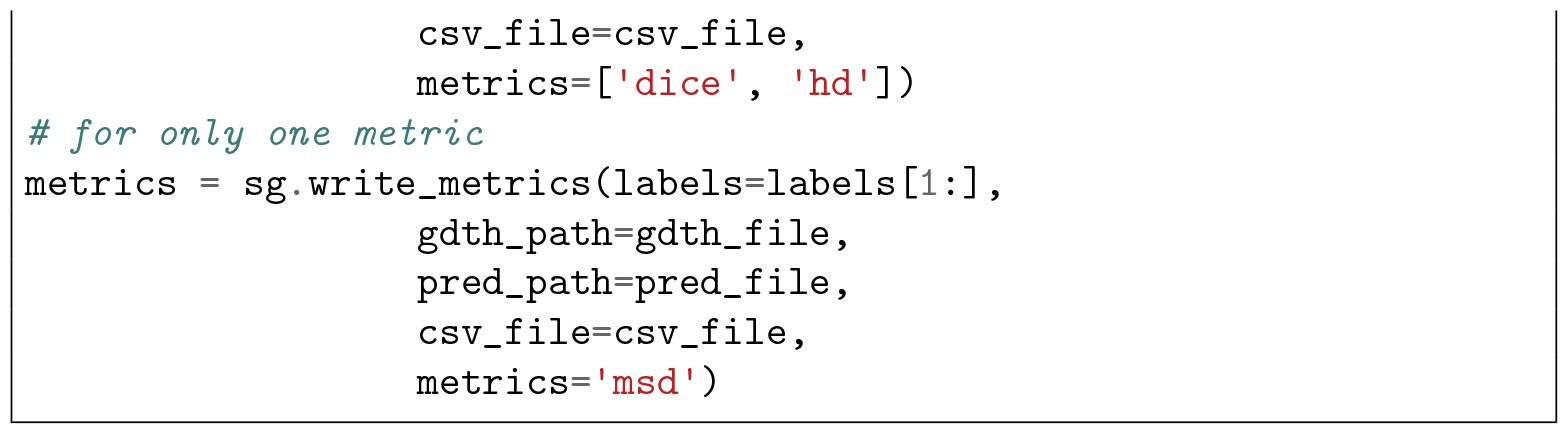
- Select specific metrics. By passing the following parameters to select specific metrics.

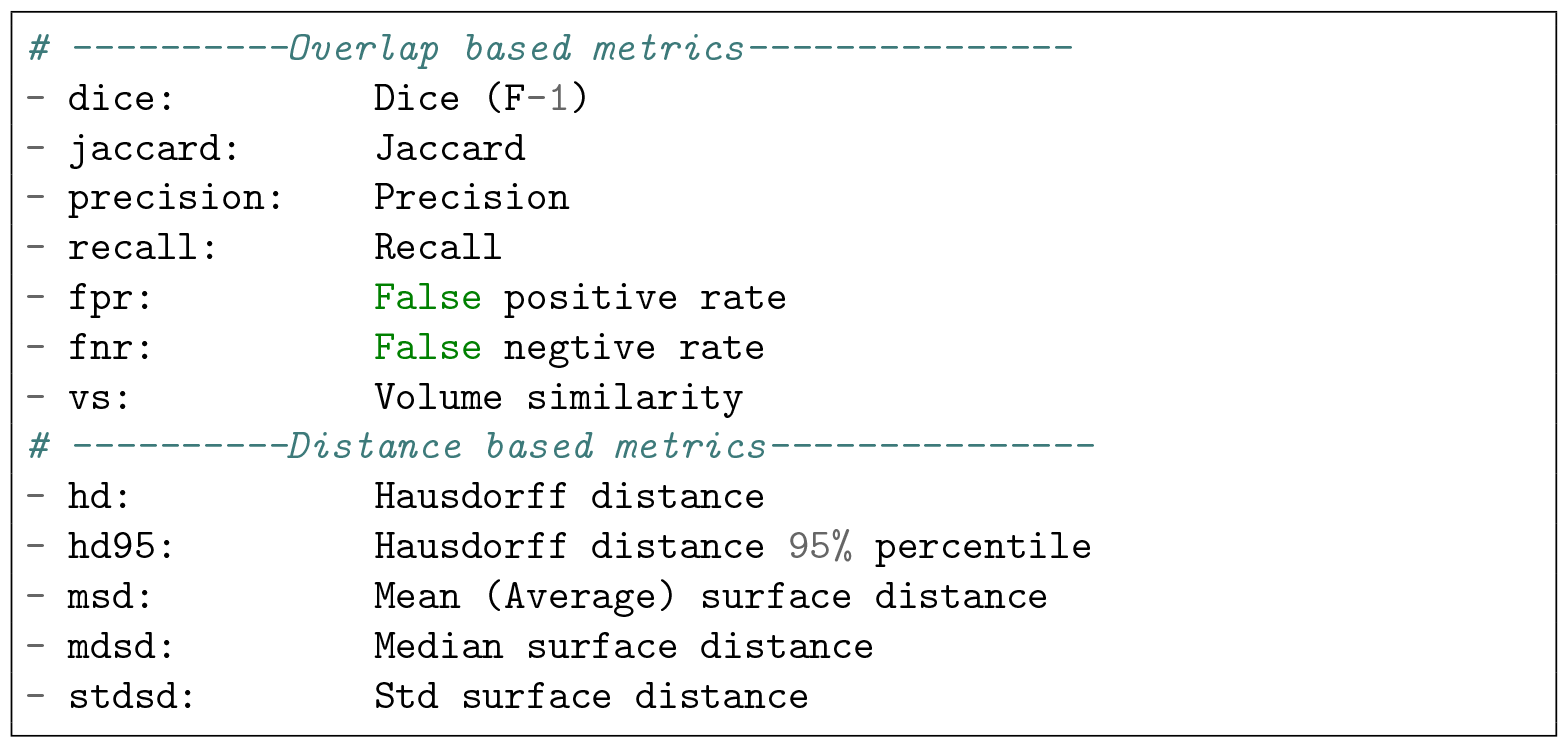 For example:

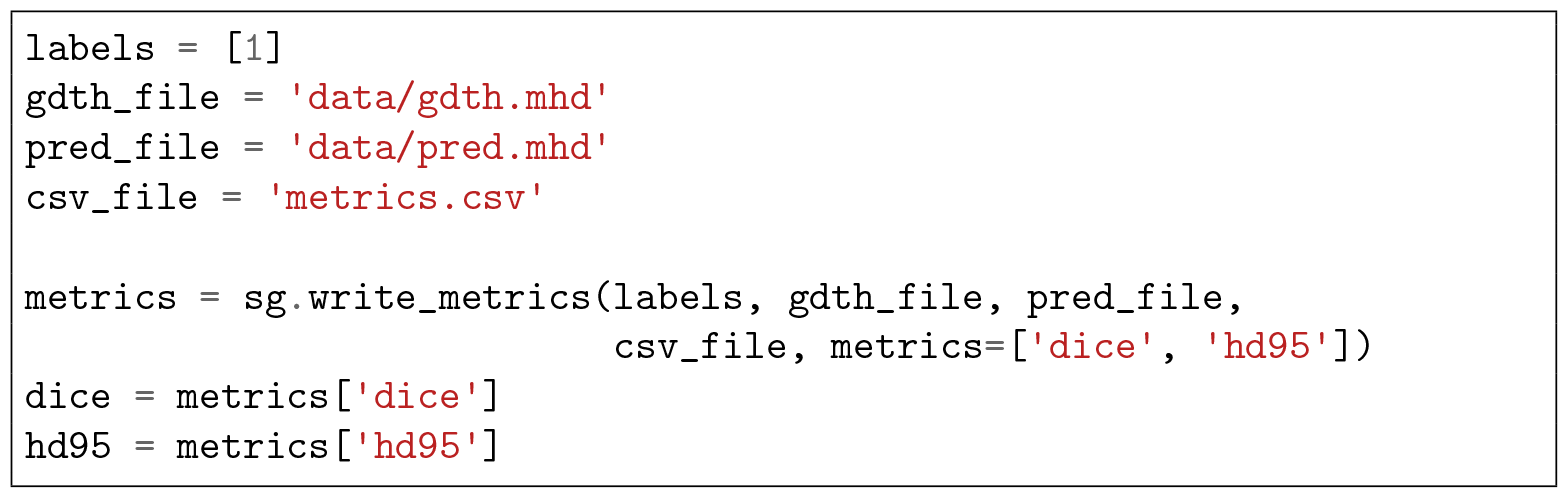

## 6 Comparison to other packages

`medpy` also provide functions to calculate metrics for medical images. Compared to it, our package `seg-metrics` has several advantages.

- **Faster**. `seg-metrics` is 5-10 times faster calculating distance based metrics (see Figure 2).
- **More convenient**. `seg-metrics` can calculate all different metrics in once in one function (shown below)

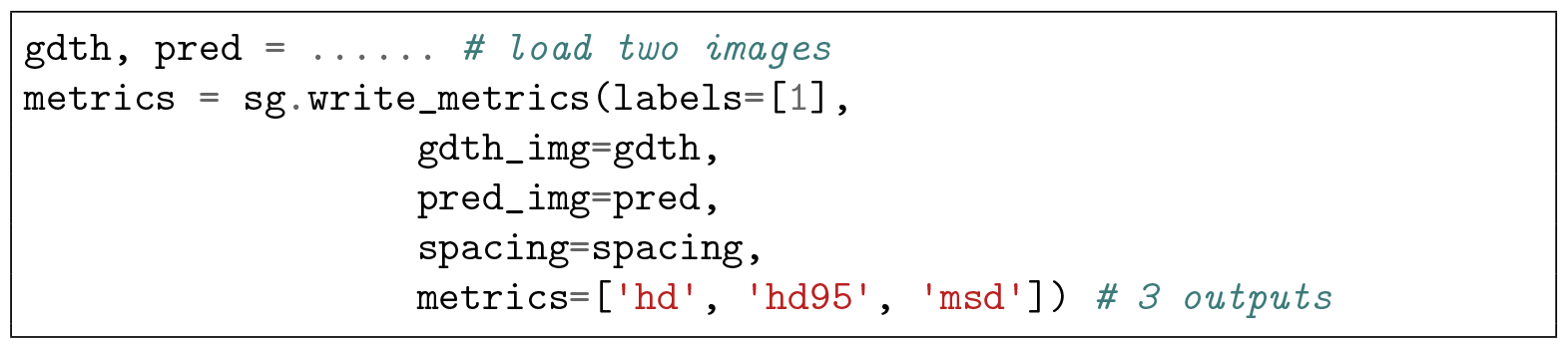

while `medpy` needs to call different functions multiple times which cost more code and time, because the calculation of each ‘hd’, ‘hd95’, and ‘msd’ will always recalculate the distance map which cost much time.

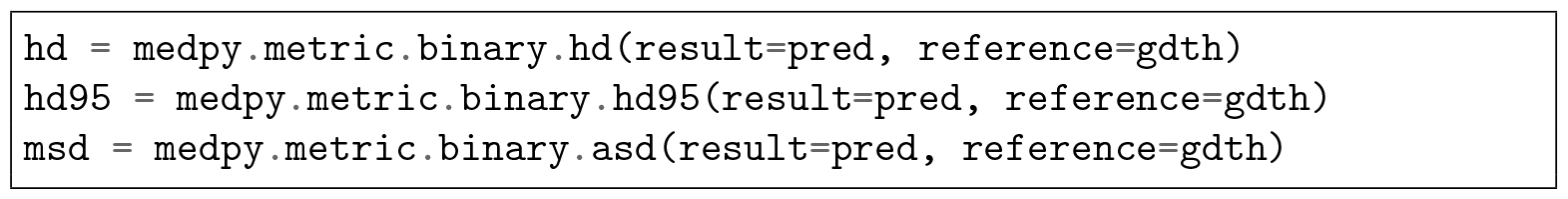
- **More Powerful**. `seg-metrics` can calculate multi-label segmentation metrics and save results to .csv file in good manner, but `medpy` only provides **binary** segmentation metrics. For instance, if there are 5 labels for an image, our `seg-metrics` can calculate 5-label metrics by one-line command while `medpy` needs to at first convert 5-label image to five binary images, then calculate binary metrics one by one,

**Figure 2:**
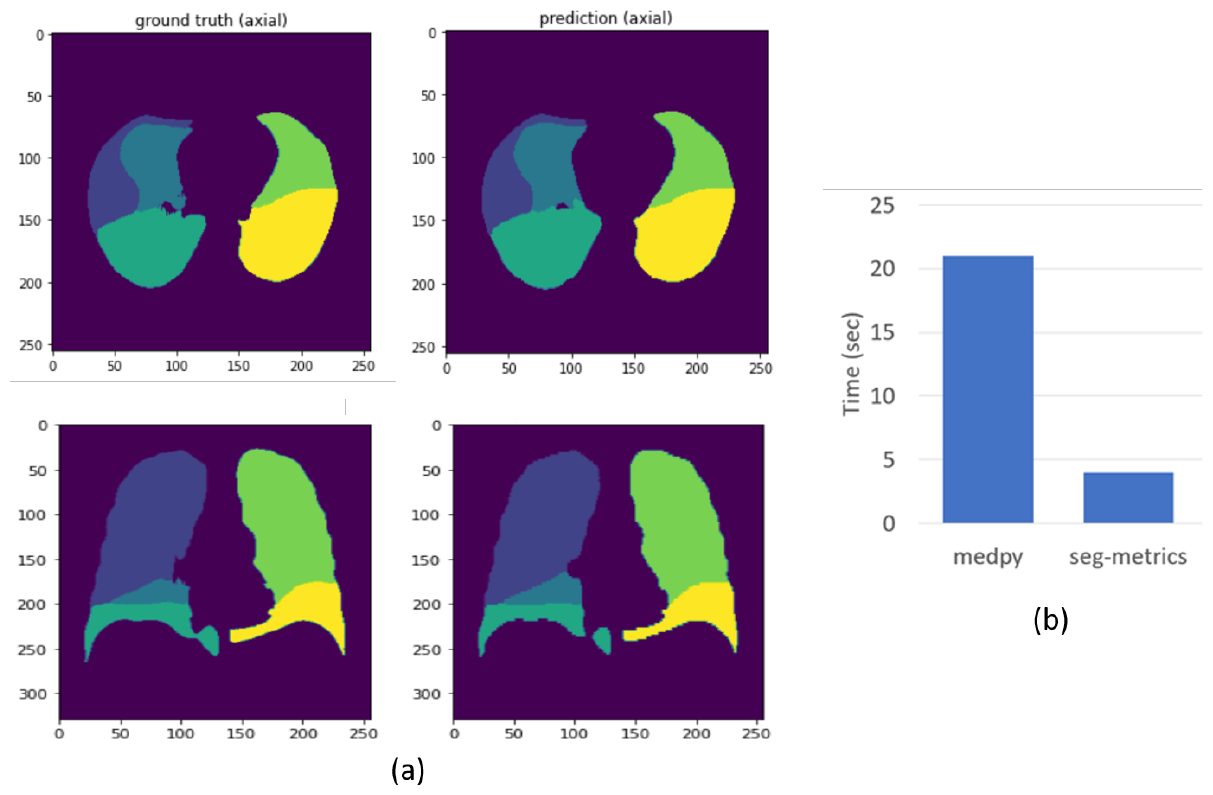
Performance comparison between `medpy` and `seg-metrics`. (a) Evaluated samples, a 3D lung lobe segmentation results (size: 256 × 256 × 256). Left: ground truth (manually annotated lobes), right: prediction (automatically predicted lobes). (b) Time comparison for the calculation of ‘hd’, ‘hd95’ and ‘msd’.

## 7 Limitation and future work

Because of time limitation for the development, there is still some space for package improvement.

- Package name. The package name is “seg-metrics” currently, as the abbreviation of “segmentation metrics”. But the dash sign “-” in the name introduced some confusion during the installing and usage of the package. Duing the installation, `pip install seg-metrics` is used. However, users need to used it by `import seg metrics`. The slight difference sometimes make new users confused and easy to make mistakes. This issue is because Python packaging system will automatically convert “_” to “-” during the installing. Because “segmetrics” has been used by other products, we may consider to change the package name to “metricseg”, “metricsrater”, “imagesegmetrics”, etc. to avoid such issue in the future.
- Supported file type. Currently, the package supports most medical image formats with suffix of `.mhd, .mha, .nii, .nii.gz, .nrrd`, etc. Because we receive some users’ requests, we will support more image formats (e.g. `.png, .jpg`) in the future.
- Usage guide. Currently, we just list the usage of different metrics, but we did not explain when to use which metrics. In the future, we hope to release a tutorial to users with some examples to which metrics are preferable in different scenarios.

## Data Availability

All data produced in the present study are available upon reasonable request to the authors

## 8 Availability and requirements

- **Project name:** seg-metrics
- **Project home page:** https://github.com/Jingnan-Jia/segmentation_metrics
- **Operating system(s):** Platform independent
- **Programming language:** Python
- **License:** MIT license
- **Any restrictions to use by non-academics:** none

## Acknowledgments

This author was supported by the China Scholarship Council No.202007720110 during the development of this package.

